# Neurofeedback-Enhanced Working Memory Training: A Proof-of-Concept for Reversing Age-Related Neural Slowing in Older Adults

**DOI:** 10.1101/2025.10.08.24316788

**Authors:** Ziming Liu, Yang Jiang, Sylvia Cerel-Suhl, Xiaopeng Zhao

**Author notes:** Correspondence should be addressed to: Ziming Liu, Ph.D. Assistant Professor School of Computer Science Gallogly College of Engineering University of Oklahoma 1170 Sarkeys Energy Center 100 Boyd St Norman, OK 73019.

## Abstract

Specific electrophysiological (EEG) brain patterns may serve as neural biomarkers of aging, predicting Mild Cognitive Impairment (MCI) in older adults, while other patterns are more closely associated with preserved brain health, such as improved working memory retrieval. In this pilot feasibility study, we developed and tested a novel visual neurofeedback (NF) protocol designed to reward EEG-based brainwave patterns linked to optimal working memory retrieval. We hypothesize that reinforcing these brainwave patterns could enhance cognitive efficiency by promoting increased alpha power, associated with focused attention, while reducing delta power, which is linked to memory lapses. Three older adults completed 8–16 NF training sessions using the protocol. Resting EEG (eyes-closed) was recorded before and after training to explore changes in brainwave patterns. Preliminary findings suggest a potential increase in alpha oscillations and a reduction in delta waves, providing initial support for the feasibility of this NF approach and motivating further evaluation in larger-scale studies.

## 1. Introduction

As individuals age, mild memory lapses often become more frequent, even in the absence of underlying pathology. Cognitive and brain aging often begins silently, up to 20 years before noticeable memory loss or other mild cognitive impairment (MCI) symptoms emerge (Caselli, Langlais et al. 2020). Importantly, age-related cognitive changes are not uniform: while certain functions decline, others may improve. For example, the efficiency of the brain’s alerting network decreases with age, whereas orienting and executive inhibitory functions may remain stable or even improve until the mid-to-late 70s (Veríssimo, Verhaeghen et al. 2022). This variability suggests that cognitive aging is not a fixed, inevitable trajectory but rather a dynamic process that can be shaped by external influences. Preventive strategies, including lifestyle changes and non-pharmaceutical interventions, are therefore crucial for promoting brain health long before the onset of MCI or dementia. Among the range of preventive strategies, cognitive training interventions have attracted particular attention.

Brain training is appealing due to its potential to directly target and improve neural activities associated with cognitive functions. Among available brain-training approaches, neurofeedback (NF) is particularly promising because it provides a closed-loop means of modulating neural circuits (Lavy, Dwolatzky et al. 2021, Trambaiolli, Cassani et al. 2021, Tseng, Tamura et al. 2021). NF has shown potential in enhancing memory and delaying cognitive decline, with studies reporting that NF-related changes in brain connectivity (e.g., measured via Functional Magnetic Resonance Imaging (fMRI)) correlate with behavioral improvements (Ramot, Kimmich et al. 2017). Additional evidence supports the use of sensory rewards, such as enhanced visual clarity, to reinforce neural circuits linked to desired behaviors like memory retrieval (Debettencourt, Turk-Browne et al. 2019). Our review of NF studies indicates a positive trend, with most reporting improvements in memory and cognition to varying extents (Jiang, Jessee et al. 2022). Despite this promise, a central challenge remains, which is how to effectively and noninvasively shape complex, cognitively relevant brain oscillations. EEG provides an accessible, noninvasive window into brain dynamics, and, critically, its millisecond temporal resolution allows for the capture of moment-to-moment fluctuations that underlie cognitive lapses (Ghassemkhani, Saroka et al. 2025). Oscillatory features (e.g., increased alpha, reduced delta (Ranasinghe, Kudo et al. 2025)) are established markers of healthier aging and MCI risk, yet protocols that leverage these features in real time as a preventive intervention for age-related decline remain underexplored. Leveraging EEG-based NF to reinforce such oscillatory patterns could therefore represent a practical pathway for large-scale, early preventive interventions.

Real-time (or near real-time) NF holds particular promise for aging populations, where timely and adaptive interventions are essential. By establishing rapid feedback loops between brain activity and behavior, such approaches can directly target the transient and early disruptions that characterize age-related cognitive decline (Chen and Ziegler 2025, Zhang, Becker et al. 2025). However, a key limitation in current clinical applications of real-time NF is the temporal and functional mismatch between neurophysiological signals and observable symptoms. Oscillatory features are often extracted through complex, high-dimensional analyses that fail to align with behavioral changes, which typically unfold within seconds (Zhang, Becker et al. 2025). This disconnect reduces the effectiveness of real-time modulation and underscores the need for paradigms where neural measures are temporally and functionally coupled with symptom-relevant states. Among potential targets, working memory (WM) is particularly well-suited due to its dynamic and short-timescale nature. This suitability stems from the fact that working memory processes unfold on a millisecond timescale and involve well-characterized neural circuits, particularly frontoparietal networks, whose dynamics can be reliably captured in real-time EEG signals (D’Esposito and Postle 2015). However, the combination of real-time neurofeedback, working memory paradigms, and EEG-based neural signals has not yet been fully developed. It remains unclear how to build a closed-loop system that can connect these elements in real time.

Beyond its suitability for real-time monitoring, WM is also clinically significant. As a core component of cognition, WM is critical for temporarily holding and manipulating information and supports related functions such as attention and cognitive control (Oberauer, Süß et al. 2000). Importantly, deficits in WM often emerge as one of the earliest signs of mild cognitive impairment (MCI), a prodromal stage of dementia. Neural activity patterns observed during WM tasks reliably predict cognitive decline and MCI risk in at-risk individuals (Li, Broster et al. 2017, Jiang, Li et al. 2021). These features position WM as an ideal target for early preventive interventions.

Building on this rationale, the present proof-of-concept study explored a novel visual neurofeedback strategy in three older adults over several weeks. The protocol integrated WM assessment, EEG-based monitoring of frontoparietal activity, and real-time feedback delivery. Specifically, participants performed WM tasks while their neural activity was recorded via EEG, and immediate feedback was provided to directly modulate WM-related processes. A central aim was to identify efficient EEG markers that are both sensitive to moment-to-moment WM fluctuations and suitable for real-time feedback.

To examine whether the protocol generated benefits beyond transient modulation, we also assessed resting-state EEG (eyes closed) before and after each training session. Resting EEG is widely recognized as a marker of brain aging and provides a trait-level index of neurophysiological change (Babiloni, Del Percio et al. 2017, Scally, Burke et al. 2018). Consistent with prior literature, **Figure 1** illustrates that increases in alpha oscillations and decreases in delta power are characteristic features of healthier aging. This allowed us to evaluate both short-term and more enduring neural effects of the intervention.

**Figure 1.**
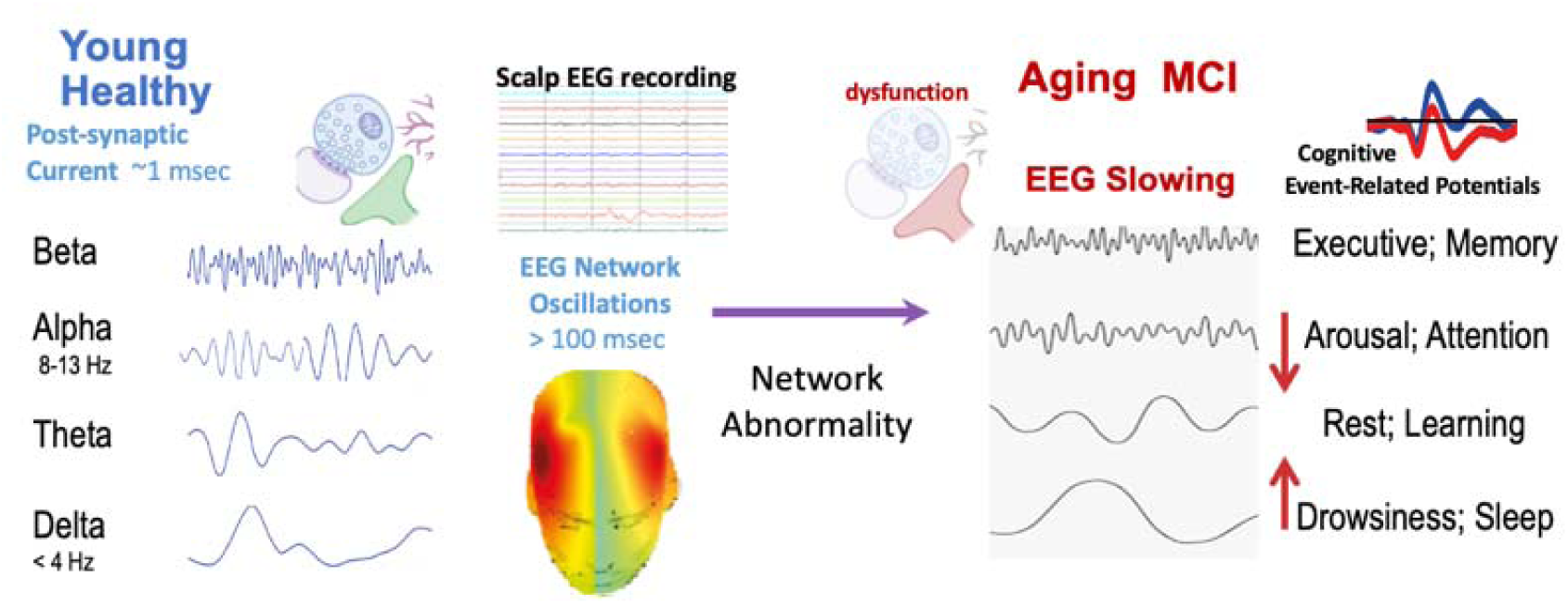
Summary of EEG slowing literature. An increase in alpha oscillations and a decrease in delta powers are indicators of brain oscillations more typical of younger and healthier brains (adapted from Jiang et al., 2024).

Prior studies that employed resting EEG as an assessment tool, rather than as part of a neurofeedback protocol, have reported that individuals with stronger working memory abilities show higher alpha activity and lower delta activity during rest (Borhani, Zhao et al. 2021, Jiang, Zhang et al. 2024). Observing similar patterns here would suggest that neurofeedback not only improves short-term working memory performance, but may also promote longer-term neurophysiological adaptations linked to healthier cognitive aging.

## 2. Methods

### 2.1 Human Participants in the Mini-Clinical Trial

We recruited four older adults from the Lexington, Kentucky VA Medical Center (VAMC). All study procedures were approved by the Institutional Review Boards of both the Lexington VAMC and the University of Kentucky. Written informed consent was obtained from all participants prior to enrollment. One participant withdrew midway due to travel constraints and surgery, leaving three male participants (aged 61-79) who completed the protocol. Two participants were left-handed, and one was right-handed.

The overall program spanned 16 weeks. The first 14 weeks were devoted to system calibration, refinement of the user interface (UI), and participant familiarization with the procedures. During this preparation phase, participants also completed structured interviews to determine an optimal session duration and ensure full comfort with the system. The final two weeks constituted the formal testing phase, conducted across five experimental sessions (Day 0, Day 3, Day 7, Day 10, Day 14). During this phase, participants engaged in neurofeedback training using novel visual stimuli (described in the *Real-Time NF Reward Protocol* section) under three reward conditions: No reward, Random reward, and NF (**Figure 2**). Sessions were held at the Sleep EEG Clinic of the Lexington VAMC. All participants had normal or corrected-to-normal vision, were not taking medications known to cause drowsiness or enhance cognitive performance and were native English speakers.

**Figure 2.**
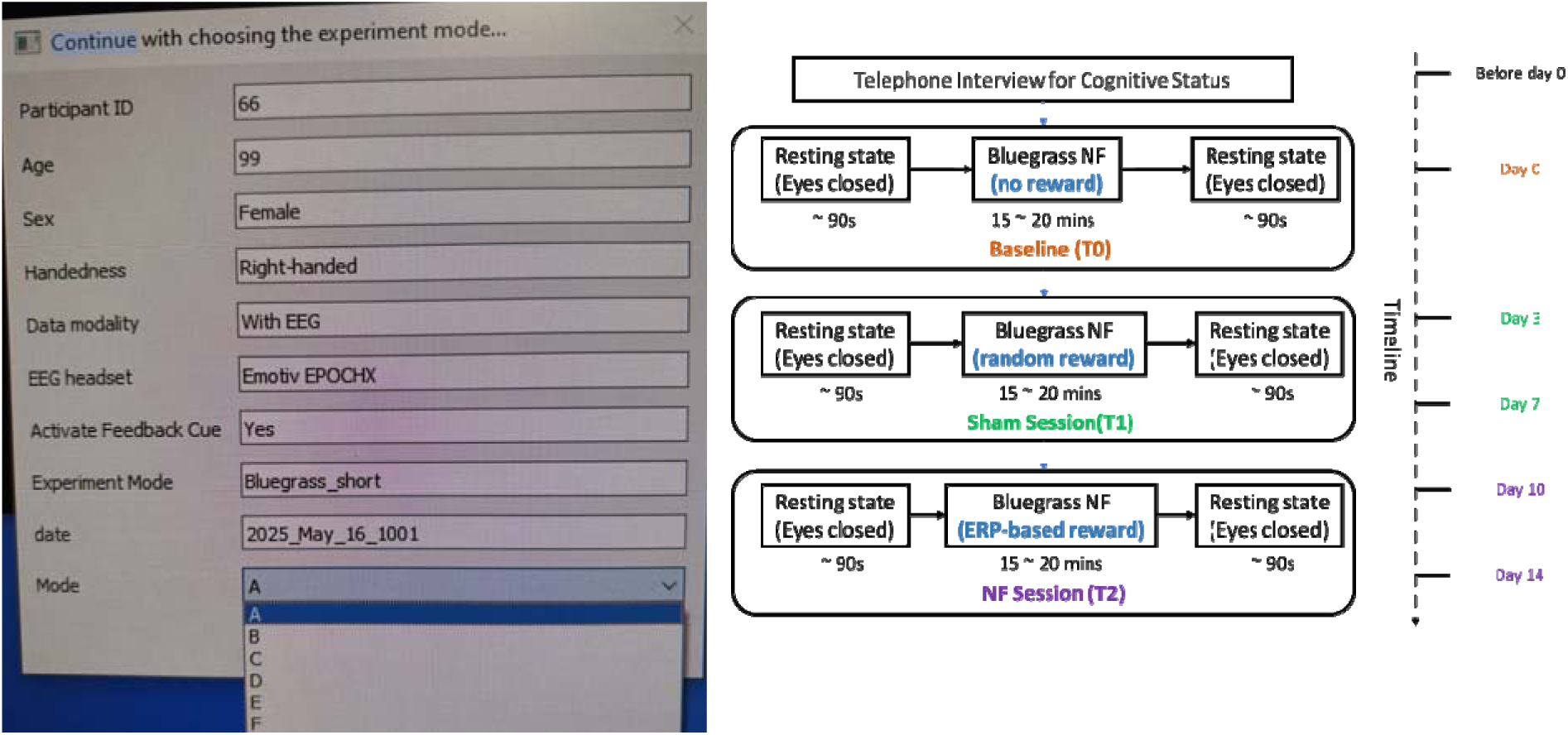
Mini clinical trial to assess NF training. <u>(NCT04446481)</u>**. 1A** (Left): The neurofeedback training was conducted at the Sleep Center Lexington VAMC. To accommodate individual differences and experimental flexibility, we also developed a customizable training interface. This interface allowed researchers to specify key parameters, including hand dominance (right-or left-hand response focus), data modality (with or without EEG), EEG headset type, feedback cue presentation (Yes, No, Sham), and experimental mode (e.g., different image sets for task variation). Note the 99-year-old female participant is shown for demonstrating purpose only and does not represent an actual human subject. This modular design supports both individualized intervention and controlled comparisons across experimental conditions **1B** (Right): Clinical trial of the neurofeedback training program to evaluate No reward, random reward, and NF conditions. The color-coded flowchart provides a clear progression of sessions over distinct days (Day 0, Day 3, Day 7, Day 10, Day 14), with each session labeled to indicate the reward type.

### 2.2 Training Timeline and Procedures

The training protocol consisted of four key sessions: an Interview session, a Baseline session (T0), a Sham session (T1), and a Neurofeedback (NF) session (T2), as depicted in **Figure 2B**. Before starting the clinical trial, participants completed a telephone-based cognitive assessment (TBCA) (Carlew, Fatima et al. 2020) to confirm that they were at a similar baseline level of memory function and exhibited no signs of noticeable memory or behavioral deficits.

On Day 0, following an initial resting-state EEG recording, participants completed two trials of the task under a no-reward condition. Although the full task structure was presented, no visual feedback or reinforcement was provided, allowing us to establish a baseline for neural and behavioral responses. On Days 3 and 7, participants engaged in the Bluegrass NF program under Sham conditions, where rewards were delivered randomly at a 25% probability, a parameter established during the 14-week preparation phase based on pilot observations. On Days 10 and 14, participants completed the NF condition, in which rewards were contingent on real-time neural performance. EEG data were collected on Day 0 (T0), Day 7 (T1), and Day 14 (T2) to track changes across conditions. Each session used a different program version to sustain engagement and mitigate practice effects.

In this study, we developed a reward protocol based on a refined version of the delayed match-to-sample task, also known as the *Bluegrass EEG Memory Paradigm* (See *Real-Time NF Reward Protocol* for details). This paradigm is recognized for its ability to assess WM capacity by requiring sustained attention from participants, making it a reliable tool for measuring individual WM retrieval and cognitive motor functions. The frontal memory-related brainwave signatures predicted MCI risk in individuals five years before diagnosis (Jiang, Li et al. 2021). The goal of the reward protocol was to encourage accurate and fast memory retrieval, reinforcing the brainwave patterns associated with optimal working memory performance.

As depicted in **Figure 2B**, participants began with a 90-second resting session, eyes closed, to capture baseline brainwave activity before transitioning to the neurofeedback training. During this period, it was imperative for participants to stay composed and attentive, ensuring they did not fall asleep. Subsequently, the neurofeedback training commenced. Upon its completion, an additional resting session was conducted to evaluate the post-training brainwave variations. An advanced experimental interface is in place, equipped with clear and user-friendly instructions, allowing participants to navigate the program independently and obviating the need for hands-on experimenter guidance. The Bluegrass resting and memory protocol takes approximately 15-20 minutes.

To provide unbiased testing of biomarkers in our longitudinal approach and reduce potential practice effects on the working memory task, we introduced eight unique test program versions (A, B, C, D, E, F, and G), each with distinct image sets for repeated sessions. To minimize any potential bias from dominant hand use affecting reaction times, participants first used the index finger of their dominant hand to indicate a memory target during the initial block of tasks. In the subsequent block, they used the index finger of their non-dominant hand to perform the same tasks. This approach ensured balanced testing of motor responses across both hands. In the first block, participants used their ***dominant hand*** to respond to target images by pressing the “A” or “L” keys on the keyboard (e.g., a right-handed participant presses “L” for a match and “A” for a non-match). After completing the first block, instructions were revisited, and participants switched to using their ***non-dominant hand*** for the second block to ensure balanced motor engagement and accuracy.

### 2.3 Bluegrass NF Reward Protocol

Our neurofeedback protocol is based entirely on EEG-derived signals and consists of two types of trials: *baseline trials* and *feedback trials*. Both trial types follow an identical working memory task structure, but differ in whether real-time visual feedback is delivered. The full procedural flow is shown in **Figure 3A**.

**Figure 3.**
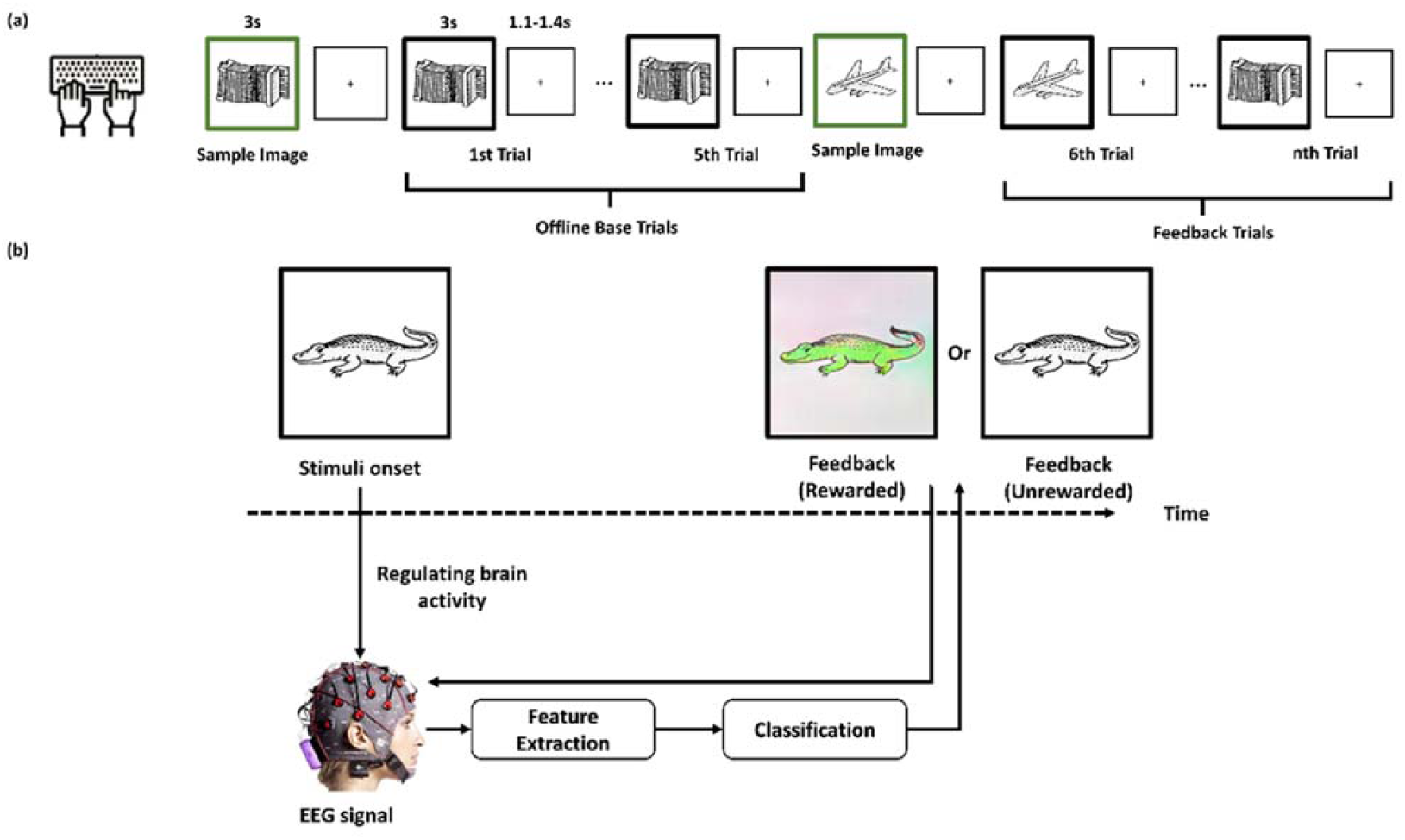
Schematic illustration of real-time EEG neurofeedback training protocol. 2A. Experiment procedure. 2B. Single trial reward (turning color) for good memory brainwaves.

#### 2.3.1 Task Structure (Baseline & Feedback Trials)

Each task block begins with a single baseline trial, followed by multiple feedback trials (typically three per block), as shown in **Figure 3A**. During the baseline trial, participants are shown two monochromatic target images for 3 seconds (encoding phase). After a variable delay (1.1–1.4 seconds), they are presented with a series of six test images, which may include targets and distractors in randomized order. Participants respond via keyboard press to indicate whether each image matches one of the targets. Importantly, all images remain in grayscale, and no neurofeedback is provided during this phase. The purpose of the baseline trial is to record EEG responses during working memory retrieval without feedback, in order to compute an individualized Event Related Potential (ERP) threshold that reflects optimal performance. In the subsequent feedback trials, participants repeat the same task using 12 test images per trial. After each image, the EEG signal is classified in real time. If the ERP amplitude exceeds a dynamically updated threshold, initially based on the baseline trial but continuously adjusted based on recent trial history (See *Processing and Reward System* for details), the image is displayed in color (reward); otherwise, it remains grayscale. This adaptive reward mechanism maintains an optimal level of task challenge and promotes sustained engagement. All images are standardized in visual format, shown on a black background (8.3 cm × 5.8 cm), and presented at a visual angle of approximately 10°. This structure is repeated across blocks and sessions.

At the beginning of each experimental session, participants completed six baseline trials of the working memory task without receiving visual feedback; all test images remained in grayscale. This phase served two purposes: to reacquaint participants with the task interface and to establish an individual ERP performance threshold for that session. The threshold was empirically set at 0.75 × the mean amplitude across correct trials during pilot testing, which provided a balance between sufficient reward frequency and task difficulty. To our knowledge, no prior studies have implemented this exact parameterization, underscoring the novelty of our approach. These values were then stored and used as the reference to guide the reward system during subsequent feedback trials, ensuring that participants had to exert sustained cognitive effort to earn visual rewards.

In the feedback phase (following baseline trials), the same working memory task is repeated; however, real-time classification of ERP features is applied after each stimulus presentation. Specifically, the EEG signal from the post-stimulus window is compared to the participant’s simultaneous ERP threshold. If the ERP amplitude exceeds this threshold, which indicates optimal working memory performance, the image is displayed in color as a form of positive feedback. If the amplitude does not reach the threshold, the image remains in grayscale. This real-time visual modulation serves as the core of the reward mechanism (**Figure 3B**), reinforcing neural patterns associated with successful memory retrieval. By providing immediate, performance-contingent feedback, this strategy aims to enhance cognitive performance and sustain participant engagement, in line with previous findings on reward-based neurofeedback (Li, Broster et al. 2017, Jiang, Li et al. 2021).

#### 2.3.2 Processing and Reward System

Throughout all trials, EEG signals are continuously recorded using the Emotiv EPOC X wireless EEG system (14 channels, 128 Hz sampling rate), following the international 10–20 system. Impedances are maintained below 10 kΩ. The system records real-time neural activity associated with working memory retrieval, particularly from frontal and parietal electrodes (e.g., F3, F4, FC5, FC6, P7, P8, O1, O2).

For preprocessing and artifact elimination in the analysis, we utilized the MNE-python library (Gramfort, Luessi et al. 2013). As shown in **Figure 4**, we focused real-time EEG signal extraction on the left frontal sites F3 and F7, which are associated with working memory processing. While WM-related P3 effects are often strongest at parietal electrodes, our prior studies using the Bluegrass paradigm with the same EPOC X system demonstrated robust group differences between MCI and healthy controls at F3 and F7 (Li, Broster et al. 2017, Jiang, Li et al. 2021). Based on this evidence and the electrode coverage of the EPOC X, we selected F3/F7 as the primary sites for real-time classification. Raw EEG data from these channels were band-pass filtered between 0.05 and 40 Hz to remove slow drifts and high-frequency noise. For each stimulus, a 3-second post-stimulus segment was extracted, and event-related potentials (ERPs) were computed within the 300–1000 ms window (termed the “post-300 epoch”). In contrast to conventional ERP averaging, which typically uses a 300–600 ms window for the P3 component, our real-time classification framework required a broader 300–1000 ms window to maximize information available from single-trial responses and to capture sustained WM-related activity beyond the initial P3 peak. Trials containing artifacts exceeding ±750 µV in amplitude were excluded from further analysis.

**Figure 4.**
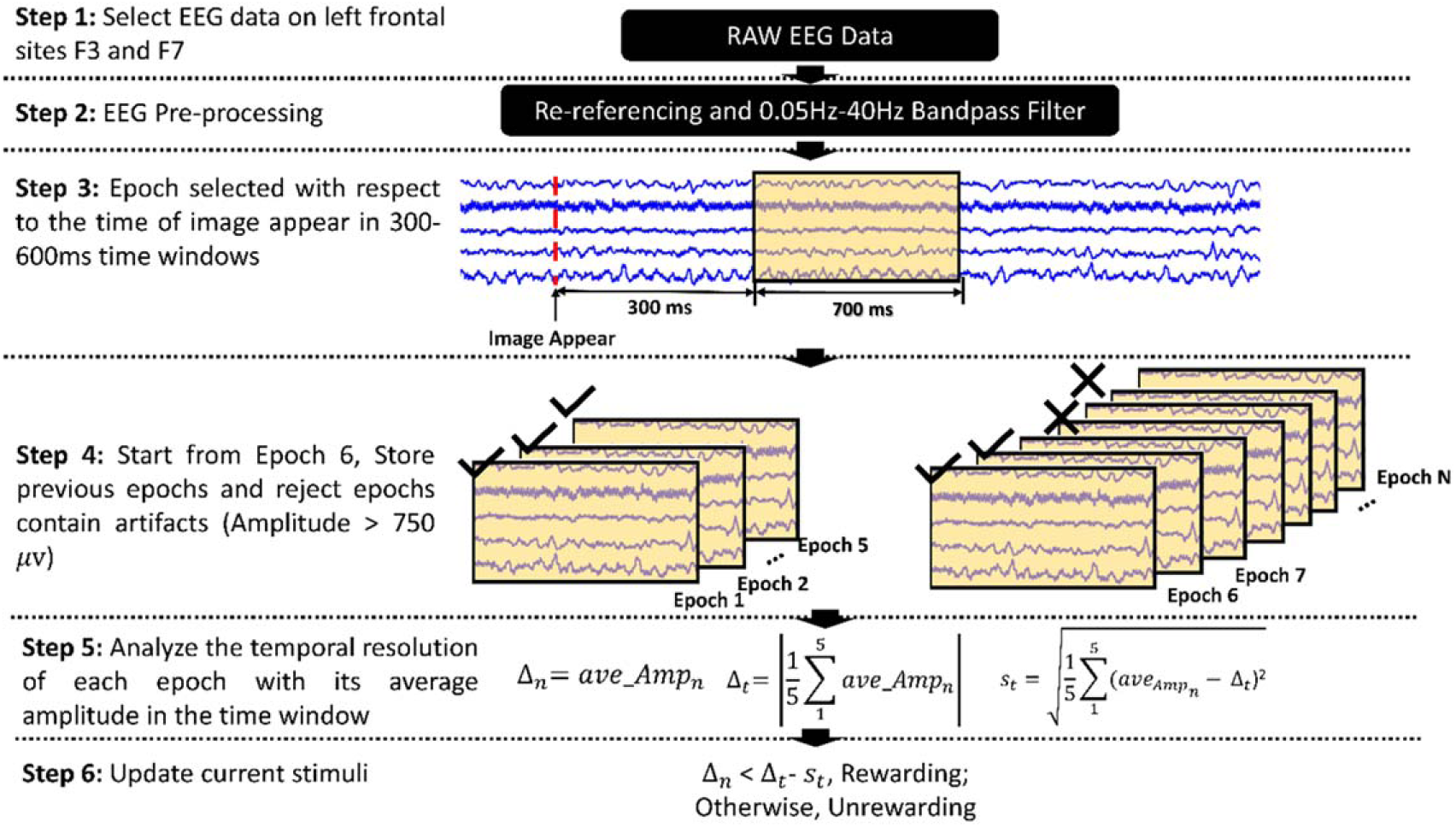
Multi-step process for extraction and classification pipeline for memory-related P3 based neurofeedback reward conditions during specific time windows.

To establish a dynamic threshold for neurofeedback classification, we computed the average ERP amplitude from the five immediately preceding stimuli within the same trial block. This moving window approach provided a continuously updated, participant-specific reference for comparison. For each new stimulus, if the ERP amplitude exceeded the average of the previous five trials, the corresponding image was displayed in color as a reward; otherwise, it remained grayscale. This real-time feedback mechanism allowed for adaptive reinforcement based on moment-to-moment fluctuations in neural performance.

### 2.4 Post EEG signal Analysis

Following the completion of all neurofeedback training sessions, we conducted offline EEG signal analysis to evaluate long-term neural changes. This post-hoc analysis is distinct from the real-time EEG activation processing used during neurofeedback training and was aimed at characterizing overall changes in spectral properties across sessions.

EEG data gathered during resting states and NF training was adjusted to the average reference and then passed through 1-Hz high-pass and 46-Hz low-pass finite impulse response (FIR) filters. Following this, to eliminate artifacts, we employed the independent component analysis (ICA) using MNE and manually removed signals that exceeded >±75 µV. ICA operates by leveraging a consistent spatial filter approach to discern maximally independent sources over time, enabling the isolation of genuine cortical EEG sources through linear disentanglement. The adopted preprocessing methodology aligns with the standards set by Wadhera & Kakkar (Wadhera and Kakkar 2021).

During the EEG data analysis for the initial resting state session, we employed a Fast Fourier Transform (FFT) on 4-second EEG segments, achieving a frequency resolution of 0.25 Hz that spanned 1–46 Hz. To mitigate the windowing effects inherent in FFT, a 4-second sliding window with a 2-second overlap was introduced. Subsequently, we measured the relative band power in the delta (d:1–4 Hz), theta (q: 4–8 Hz), alpha (a: 8–13 Hz), beta (b: 13–28 Hz), and gamma (g: 28–46 Hz) frequency bands across all 14 EEG channels. This analysis facilitated the extraction of the absolute power for each frequency band and enabled the computation of the magnitude-squared coherence amongst channels. Our objective is to discern variations in the average relative band powers across three distinct sessions (T0, T1, and T2) for the participants.

During the NF training, a 3-second time window was initiated from each stimulus onset for EEG data analysis. We also computed the relative band power in the delta, alpha, beta, and gamma frequency bands. To discern the variations in relative band powers between the Sham and NF training sessions, we performed a t-test. This analysis was aimed at assessing the significance of differences between the two sessions for each frequency band.

### 2.5 Evaluating NF Effectiveness

To assess the effectiveness of this novel reward protocol, participants engaged in neurofeedback training. The training began with a baseline trial, followed by ten feedback trials where reward criteria were based on the baseline-defined threshold. The task was divided into two blocks, each with three trials.

## 3. Results

In this clinical trial involving longitudinal NF training, we examined the relationship between EEG indicators of memory performance and training intensity over three sessions. We analyzed the reaction times (in seconds) and accuracy scores of the three participants throughout these sessions. Additionally, we investigated the correlation between neural measures and behavioral outcomes from the clinical trials.

### 3.1 Behavioral Results

To assess participants’ behavioral changes across sessions, we focused on two main markers: the accuracy of their responses (correct vs. incorrect) and their reaction times. Evaluations were scheduled at three time points (**Figure 2B**) to maintain consistent intervals: Day 0 (T0), Day 7 (T1), and Day 14 (T2).

*Accuracy.* Initially, we evaluated the accuracy of participants’ responses. However, most trials resulted in correct responses across participants, limiting the variability in accuracy scores. As a result, we focused on the counts of **correct** and **incorrect** responses under different conditions (session vs. target or non-target stimuli) to represent accuracy. **Table 1** summarizes the accuracy scores across three participants for the three sessions.

**Table 1.**
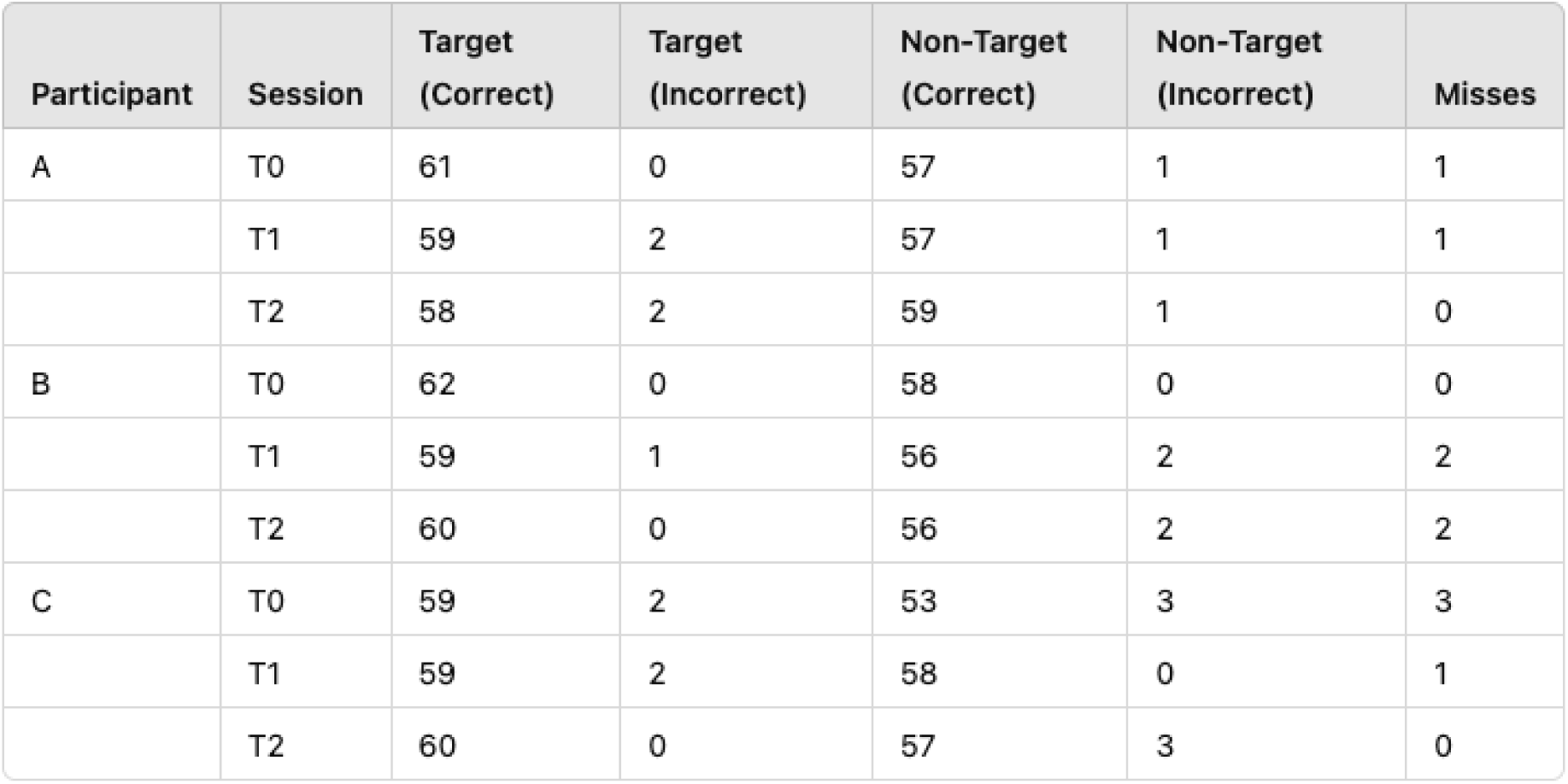
Behavioral percent (%) accuracy across participants in three training sessions for target and non-target visual stimuli.

| Participant | Session | Target (Correct) | Target (Incorrect) | Non-Target (Correct) | Non-Target (Incorrect) | Misses |
| --- | --- | --- | --- | --- | --- | --- |
| A | T0 | 61 | 0 | 57 | 1 | 1 |
|  | T1 | 59 | 2 | 57 | 1 | 1 |
|  | T2 | 58 | 2 | 59 | 1 | 0 |
| B | T0 | 62 | 0 | 58 | 0 | 0 |
|  | T1 | 59 | 1 | 56 | 2 | 2 |
|  | T2 | 60 | 0 | 56 | 2 | 2 |
| C | T0 | 59 | 2 | 53 | 3 | 3 |
|  | T1 | 59 | 2 | 58 | 0 | 1 |
|  | T2 | 60 | 0 | 57 | 3 | 0 |

The accuracy data show that all participants correctly identified the stimuli across sessions. There were no significant differences observed between target match and non-target match conditions or across the T0, T1, and T2 sessions.

*Reaction Times.* Next, we evaluated the participants’ reaction times during each session.

**Table 2** presents the average reaction times across the three sessions. Participants A and B demonstrated consistent reaction times (∼0.7-0.8 seconds), while Participant C exhibited slightly longer reaction times, which could be attributed to age and cognitive status. A one-way ANOVA was performed to evaluate the effect of training sessions on reaction time, and the results (F = 0.66, *p* = 0.82) indicate no significant differences in reaction times across sessions.

**Table 2.** Average reaction times (with standard deviations) across participants in three training sessions.

| Participant | Session | Reaction Time (s) |  |
| --- | --- | --- | --- |
|  |  | Mean (msec) | Std |
| A | T0 | 800 | 280 |
|  | T1 | 860 | 310 |
|  | T2 | 840 | 250 |
| **B** | **T0** | 780 | 240 |
|  | **T1** | 890 | 320 |
|  | **T2** | 730 | 150 |
| **C** | **T0** | 1130 | 410 |
|  | **T1** | 1030 | 350 |
|  | **T2** | 990 | 320 |

#### 3.1.1 Rewarding brainwaves during working memory retrieval

To further investigate the effect of neurofeedback on working memory capacity, we analyzed participants’ performance in reward versus no-reward conditions. In the reward condition, images turned color 25% of the time as a form of feedback. Behavioral analyses focused on accuracy and reaction time under these two scenarios.

In this context, “reward” means that the images turn to color based on the real-time threshold we designed, while “no-reward” means the images remain black-and-white. Specifically, T1 (sham) randomly shows reward or no-reward, whereas T2 (NF) trials reward or no-reward based on real-time brainwave activity.

This design is intended to reinforce brainwave patterns that are indicative of successful memory retrieval, as measured by ERP amplitudes exceeding a dynamic threshold during the retrieval phase. Based on baseline brainwave observations, we set the threshold to around 25% to trigger the images to turn to color. Consequently, **Table 3** shows that only 25% of the trials are marked as correct and colored, while 75% are correct but remain uncolored.

**Table 3.**
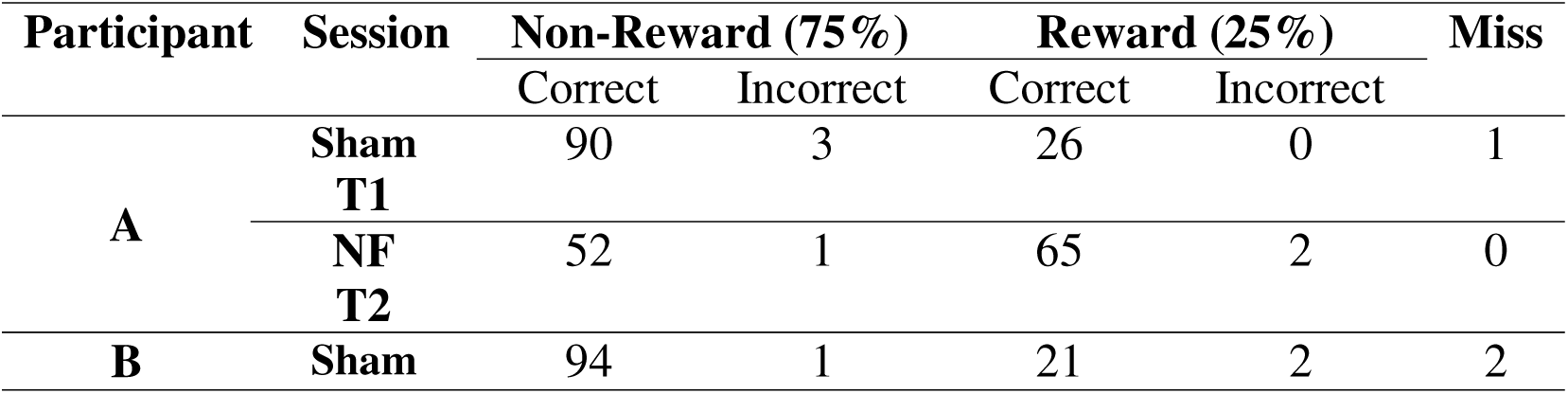
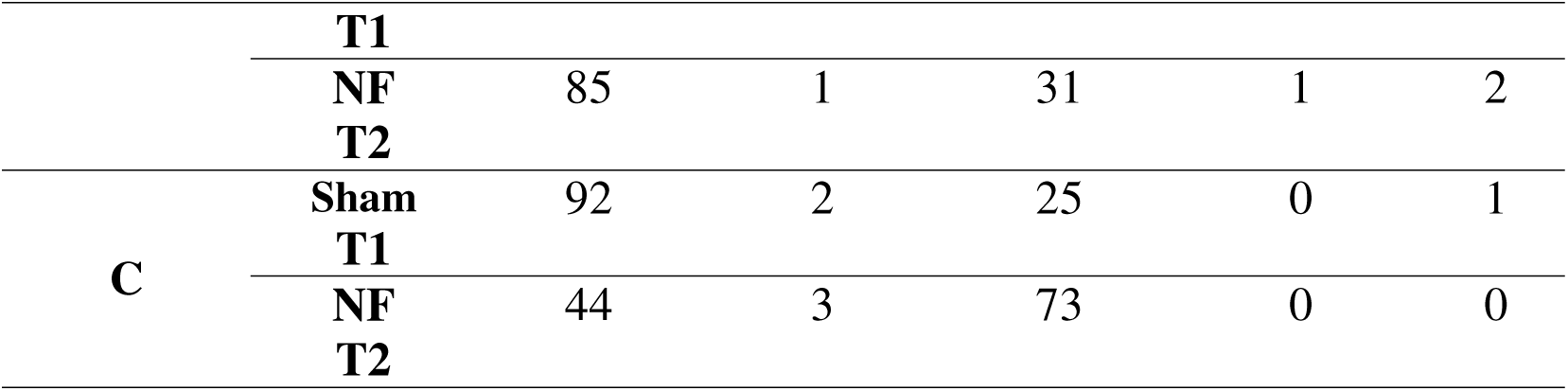
Accuracy scores in Reward vs. Non-reward conditions.

**Table 3** summarizes the accuracy scores, revealing that Participants A and C showed improvements in the reward trials compared to the sham session. However, Participant B exhibited no noticeable difference in performance between reward and sham conditions. Post training interview revealed that Participant B was trying hard to figure out or “beat” the program, which was not part of the training.

The 25% reward rate in the sham condition was deliberately selected to reflect the approximate rate of color changes observed during the baseline ERP calibration phase. This was intended to ensure that the sham and NF sessions were visually comparable, while only the NF session provided meaningful, brainwave-contingent feedback. However, because sham feedback was randomly assigned and unrelated to actual brain activity, it may have introduced perceptual noise or distraction, thereby undermining performance for some participants. By contrast, neurofeedback delivered in the NF condition reinforced brainwave patterns associated with successful memory retrieval, potentially enhancing accuracy and engagement. The discrepancy between Participants A/C and Participant B further highlights the importance of user strategy: Participant B’s effort to interpret the system may have inadvertently disrupted the implicit learning mechanism, reducing the effectiveness of neurofeedback.

The reaction times during NF and reward (T2) conditions are faster than those during sham and *random* reward conditions (grey shade), where the color change has no relation to good brainwaves (**Table 4**). There is no consistent pattern when comparing the trials of reward versus no-reward within the sham or NF conditions. This suggests that the protocol did not significantly affect reaction time, although it did improve accuracy for some participants.

**Table 4.** Reaction times (seconds) in reward vs. sham conditions.

| Participant | Session | Reaction Time (s) |  |  |  |
| --- | --- | --- | --- | --- | --- |
|  |  | Reward |  | Non-Reward |  |
|  |  | Mean | Std | Mean | Std |
| A | T1 | 0.88 | 0.34 | 0.76 | 0.14 |
|  | T2 | 0.82 | 0.19 | 0.84 | 0.29 |
| B | T1 | 0.88 | 0.31 | 0.92 | 0.35 |
|  | T2 | 0.71 | 0.14 | 0.78 | 0.18 |
| C | T1 | 1.02 | 0.35 | 1.10 | 0.35 |
|  | T2 | 0.95 | 0.25 | 1.01 | 0.36 |

### 3.2 EEG Results during Neurofeedback Training

To assess differences in EEG frequency bands between the sham (T1) and neurofeedback (T2) sessions during the Bluegrass NF training, we conducted ANOVA tests. These tests compared variations across five frequency bands under different conditions: target match vs. non-target and rewarded vs. unrewarded trials.

The results for sessions T1 and T2 are presented in **Figure 5**, which provides a detailed topographic comparison of EEG data across different frequency bands (δ, θ, α, β, γ) and task conditions (Non-target Unrewarded, Non-target Rewarded, Target Unrewarded, Target Rewarded). Significant differences (p < 0.05) were observed in delta and alpha band oscillations, particularly in the left hemisphere. This layout highlights the ANOVA results, visually represented by p-value heatmaps, with a color scale ranging from green (high p-values, indicating less significance) to red (low p-values, indicating higher significance). The use of distinct frequency bands and conditions offers insights into how different neural oscillations respond to reward and target-related factors. Notably, rewarded trials displayed fewer regions with significant differences compared to unrewarded trials, indicating more stability in EEG activity during reward-based training.

**Figure 5:**
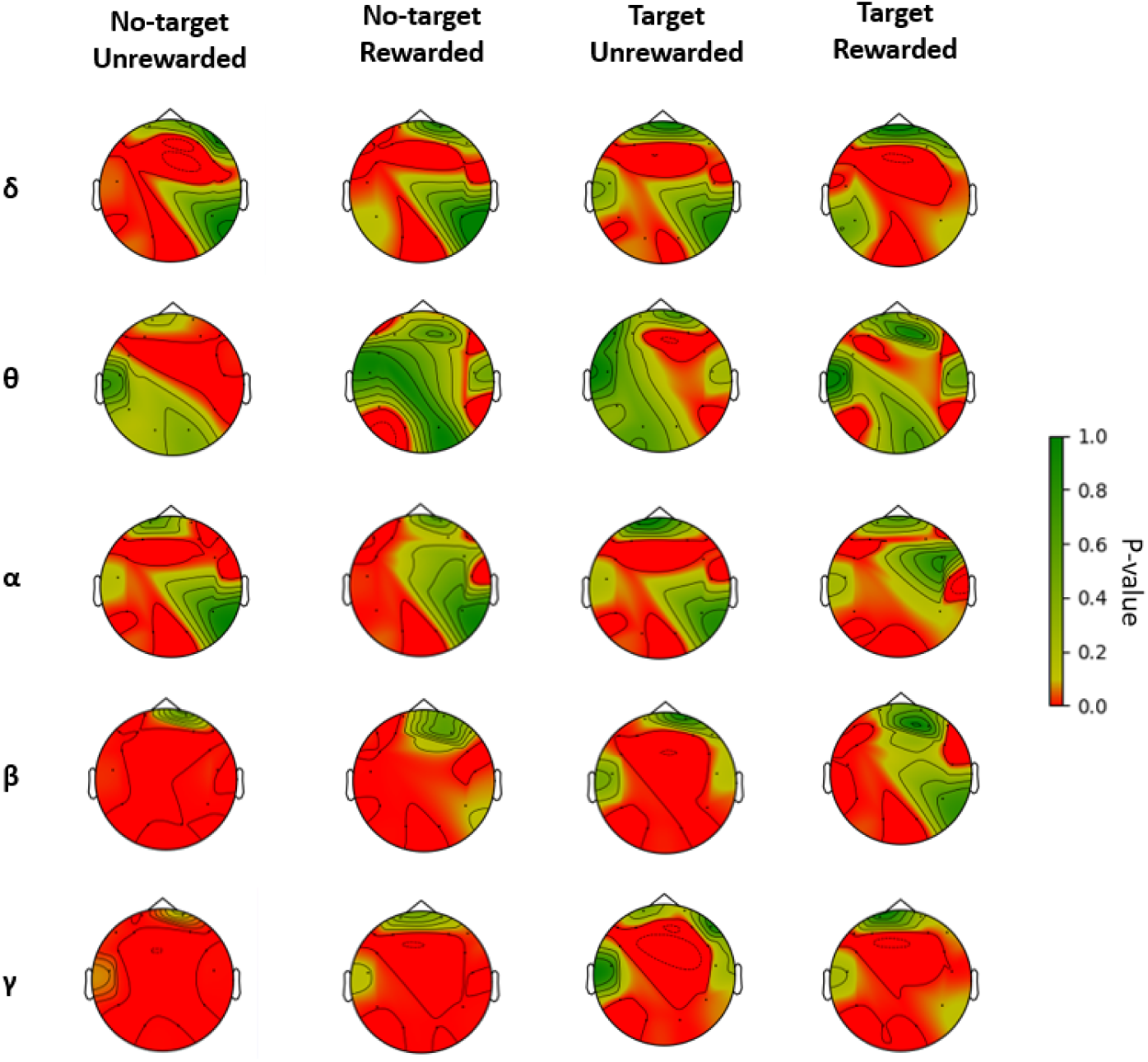
Topographic representation of ANOVA results comparing average frequency band powers between the No reward Sham (T1) and Reward NF training (T2) sessions and for participants’ EEG data during the Bluegrass NF training.

Specifically, delta (δ) band exhibits more red areas (lower p-values) in Target rewarded than target unrewarded conditions, suggesting significant differences in activity when rewards are applied. The theta (θ) band shows moderate variations across conditions, with lower p-values observed in the unrewarded nontarget trials, which is often associated with visual attention and memory processes. The alpha (α) band highlights more different activity between conditions indicating reward-related changes in this frequency. Finally, both beta (β) and gamma (γ) bands display strong differences in p-values across sham conditions. The higher frequency β band differ significantly in target reward and no-reward manipulations in this NF training protocol.

### 3.3 Assessing Effectiveness of Memory-related NF Training

To ensure the quality of our EEG data, we examined the average relative frequency bands across three participants over three training sessions: T0, T1, and T2. **Figure 6A** illustrates that, compared to the Baseline and Sham sessions, the NF training session enhanced the intrinsic brain oscillations in the alpha bands of the occipital region (O1 and O2 electrodes). This observation is consistent with prior research indicating a link between these oscillations and symptoms associated with the studied condition (Borhani, Zhao et al. 2021). We also observed a similar decrease in the delta band in the left parieto-occipital region (P7 electrode). These findings align with previous studies related to biomarkers associated with cognitive improvement (Babiloni, Del Percio et al. 2015). Additionally, when compared to the baseline, the Sham session also exhibited an improvement in the alpha band in the occipital region.

**Figure 6:**
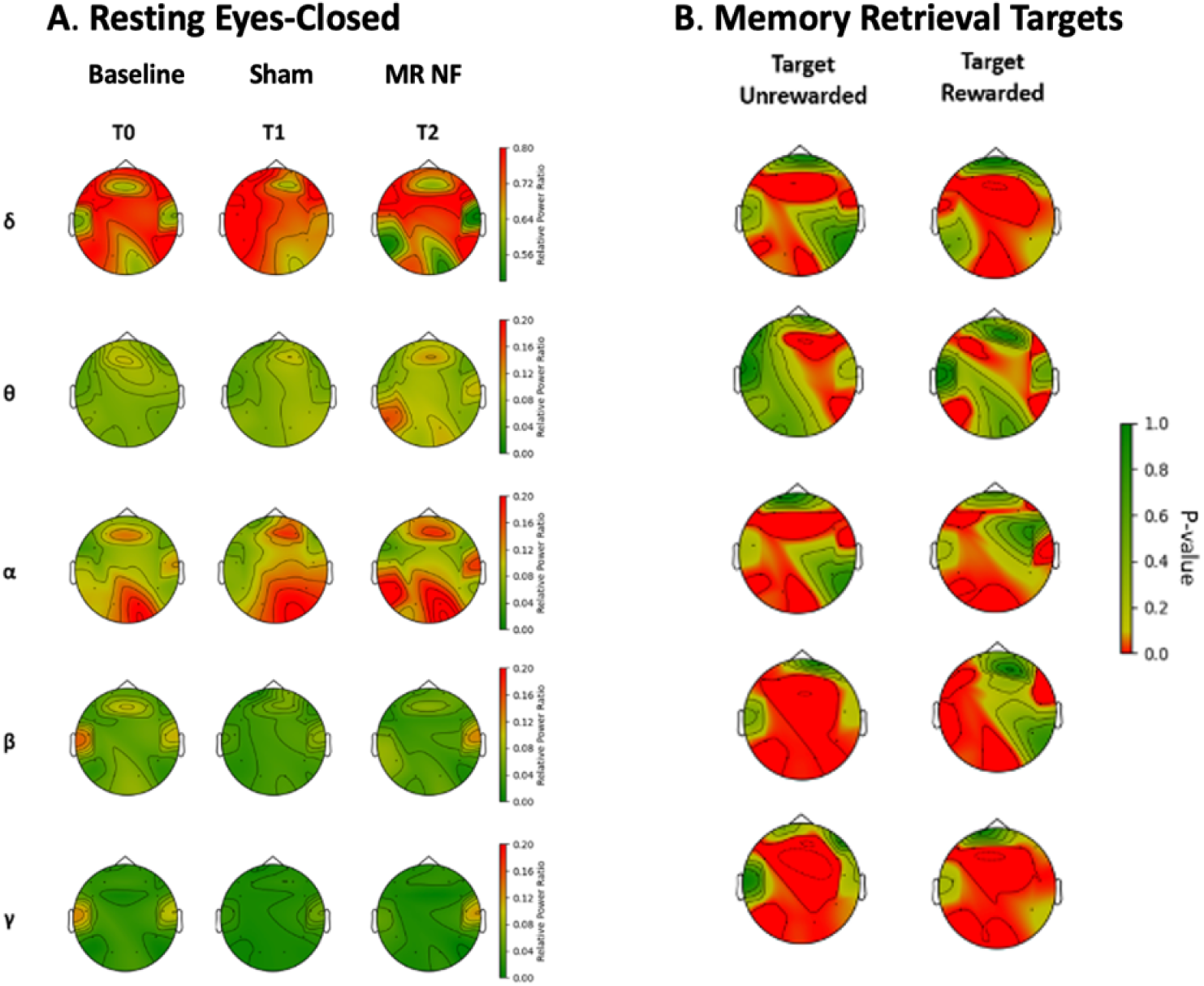
(A) Topographic plot of average frequency band powers through participant under three training session (T0, T1, T2). (B) Similar patterns are observed during the memory-related NF training.

The eyes-closed resting EEG serving as neural biomarkers for brain functions were compared. The increased alpha and reduced delta are in the direction of younger brain patterns, both Sham and NF have shown changes in the right direction. **Figure 7** shows a male participant in the 75–79 age range whose alpha and delta frequencies moved in the right direction of younger brain within 10 days.

**Figure 7:**
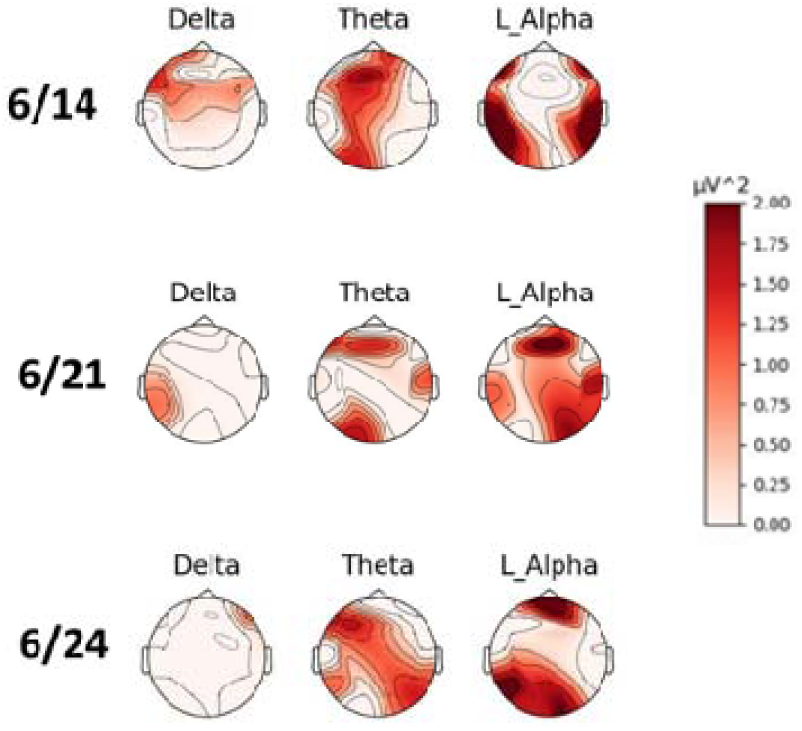
The frequency power of delta, theta, and lower alpha during resting eyes-closed EEG before each of th three sessions of memory-related NF training in a male participant in the 75-80 age range in June 2022. The change of intrinsic brain activity is in the direction of younger brain patterns 10 days later on 6/24.

Upon examining the topographic plots of relative frequency band powers across three participants, we observed that, apart from Participant B (**Figure 8**), the other two participants exhibited patterns consistent with the average. Following each rehabilitation session, discussions were held with each participant to gauge their feedback on the program. Notably, Participant B disclosed intentionally employing alternative strategies, such as deliberately avoiding memorizing the stimuli to test how the images trigger color perception. This deliberate deviation in approach is the reason why Participant B’s data stands out as an anomaly. Alternatively, Participant B is in age range 61-65, much younger than the two adults in age range 76-80 who may benefit more from the training.

**Figure 8.**
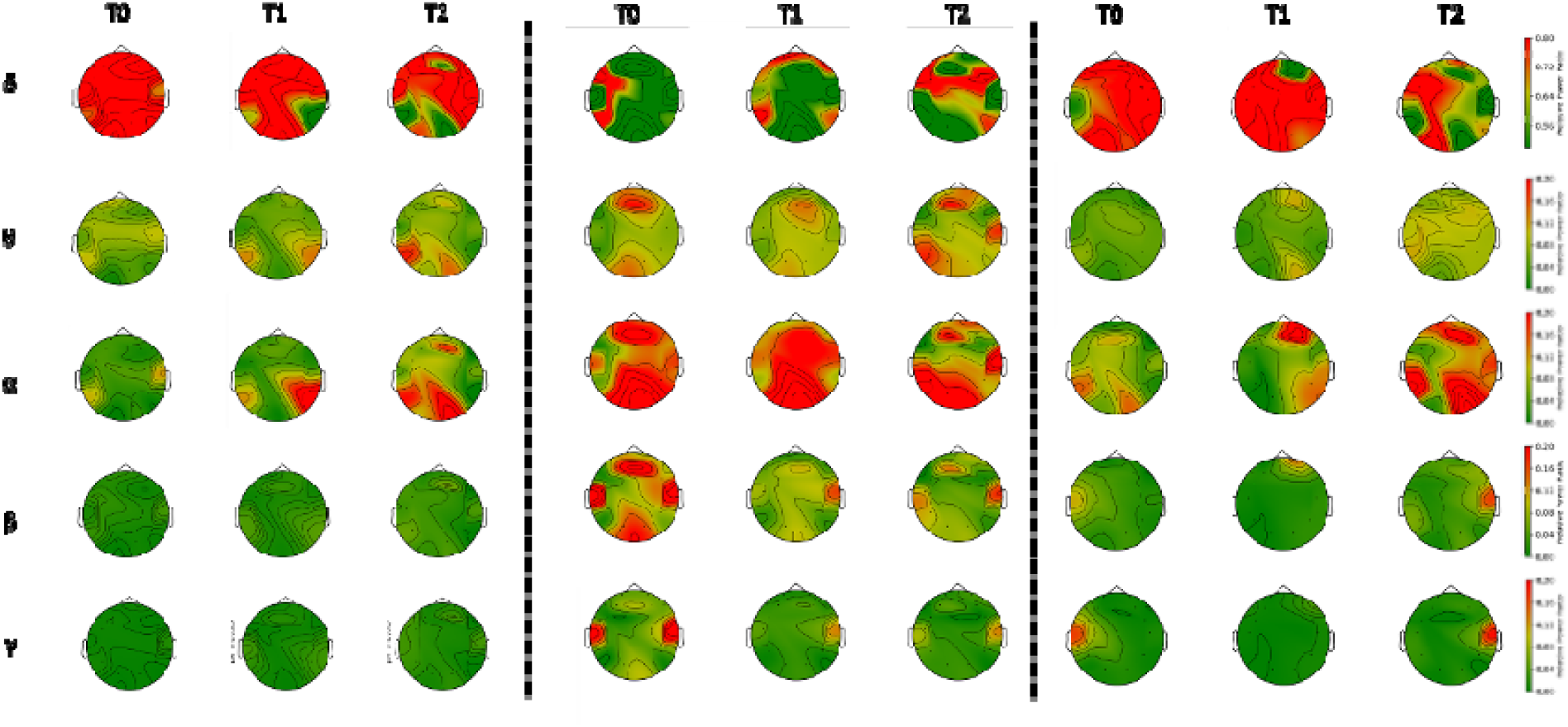
Topographic plot of relative frequency band powers across three participants under three training sessions (T0, T1, T2). Participant A (age range 75-79, left), Participant B (age range 60-64, middle), and Participant C (age range 75-79, right).

## 4. Discussions

### 4.1 Summary of Results – Individualized NF Training the Younger Brain Direction

In response to the need for targeted interventions, we developed **Bluegrass Neurofeedback**, a personalized protocol designed to enhance working-memory–related brain activity in older adults. This mini-clinical trial involved three participants, combining in-person EEG training sessions with remote cognitive assessments. The protocol used neuro-signal– inspired algorithms to reward brainwave patterns linked to optimal WM performance in frontal regions. EEG oscillations are well-established biomarkers of dementia risk, as changes during both resting and task states can predict transitions from MCI to dementia (Jiang, Jessee et al. 2022).

Of the three participants, two older adults (age range 76-80) showed directional improvements in intrinsic brain signals toward “younger” patterns within 10 days, whereas the younger participant (age 60-65) showed no clear benefit after eight weeks of twice-weekly training. Exploratory analyses indicated modulation of delta and alpha oscillations, with NF (rewarding memory-related brainwaves) producing more consistent changes than sham (random reward). Both NF and sham sessions trended toward increased alpha and reduced delta, patterns associated with healthier brain function. Together with prior literature (Li, Broster et al. 2017, Jiang, Li et al. 2021), these pilot findings suggest that NF may be a promising component of combined interventions to support memory in aging.

### 4.2 Reversing “Neural slowing”

Early neural biomarkers of dementia risk, such as subtle changes in neurosynaptic activity, can be detected by EEG before major neuronal damage or structural brain changes occur(Zheng, Zhao et al. 2023). One key indicator of early AD risk is synaptic dysfunction, which often manifests as network synchronization issues, including hyperactivity in both subcortical and cortical networks.

The physiological model of intrinsic EEG oscillations in normal cognition versus Alzheimer’s disease shows a clear shift also known as “EEG slowing”: slowing of intrinsic rhythms reflects disconnection in the brain’s networks. This disconnection is indicated by EEG biomarkers, where reduced alpha activity is observed in AD compared to NC, increased delta power and elevated resting-state theta (Ranasinghe, Kudo et al. 2022). Notably, in our previous study involving 43 cognitively normal older adults from University of Kentucky-ADRC, some individuals exhibited AD-like increases in delta activity, further highlighting the potential of EEG as a tool for early detection (Borhani, Zhao et al. 2021). By contrast, increases in task-evoked frontal-midline theta have emerged as biomarkers of proactive control, but not reactive control, aligning with our own research(Eschmann and Mecklinger 2022).

In the present study, two Veterans in age range 75-80 who completed three NF sessions within a two-week period showed improvements in resting EEG oscillations, including reduced delta activity and increased low alpha. Higher frequencies such as beta and gamma (not shown), typically linked to cognitive-motor functions, showed stable patterns without systematic increases or decreases across sessions. During task performance, both target match and non-match retrievals exhibited similar oscillatory patterns, with rewarded trials producing more spatially focused modulation compared to unrewarded trials. This aligns with decades of neuroimaging evidence showing that younger brains recruit focal networks more efficiently, whereas older brains rely on more diffuse activity to achieve comparable performance.

### 4.3 Limitations and Future Combined Attention and Working Memory Training

In this study, we conducted a preliminary investigation to assess the viability of using WM-based NF for real-time brain training. The results indicate cognitive improvement in two out of three participants, suggesting potential efficacy. However, due to the limited sample size, it is premature to draw definitive conclusions. Thus, the primary limitation identified is the small dataset. Future efforts will aim to recruit a larger cohort to ascertain the method’s effectiveness across a broader population. Additionally, we observed that one participant (B) who did not benefit from training attempted to decipher the neurofeedback strategy, resulting in atypical measurements compared to the others. To address this in future studies, improved instructions will be provided to ensure participants can easily follow the protocol, thereby enhancing data accuracy.

A challenge identified in the current study is the benchmarking of rewards. Without baseline data, a fixed reward number was used, limiting the personalization of feedback. To address this, we plan to implement a data-driven approach with a larger dataset in future studies. This will allow us to develop dynamic benchmarks for reward administration, enabling feedback to be more effectively tailored to individual performance levels.

Our present results show that both target match and non-match retrievals exhibited similar patterns. Notably, rewarded trials displayed focused brain regions with significant differences compared to unrewarded ones, which is consistent with decades of neuroimaging results in cognitive aging showing that younger brains are more efficient and focused, while older brains engage more diffused regions to accomplish a cognitive task. These findings give us more confidence in developing the WM-NF program.

A longitudinal study tracking healthy older adults over a decade highlighted the predictive power of differential brainwave responses between working memory targets and non-targets across three left frontal electrodes (Jiang, Li et al. 2021). These findings underscore the potential of leveraging real-time ERP features from these electrodes as biomarkers for neurofeedback delivery decisions. Moreover, a recent study employed functional near-infrared spectroscopy (fNIRS), a more accurate yet efficient technique, in memory-related neurofeedback training. The findings further highlight the importance of the prefrontal cortex in memory enhancement(Yang and Wang 2025).

In therapeutic NF, particularly those targeting cognitive control, the integration of feedback information, the establishment of intrinsic rewards, and the facilitation of behavioral self-regulation are crucial. These processes are intimately linked with attention—a multifaceted concept related to the distribution of working memory across neural representations in the brain. Attention plays a critical role in allocating brain resources to working memory (Knudsen 2007), while high-level cognition relies on working memory to learn tasks (Cowan, Elliott et al. 2005, Gazzaley and Nobre 2012, Hsueh, Chen et al. 2016, Fan, Che et al. 2024). The shared neural mechanisms support the theory of a mutual connection between these two brain functions. Thus, event-related (memory and/or attention) brainwave patterns should be excellent and focused targets of NF training. The reason it has not been widely used in brain training is partially due to the difficulty of real-time implementation and complexity of attention during encoding, maintenance, and retrieval of items held in working memory.

As the aging population grows rapidly, these proposed non-pharmacological interventions to boost brain functions in older individuals will meet the urgent and unmet need to reduce the risk of cognitive decline in asymptomatic aging individuals. This cutting-edge rehabilitative research promises more impactful studies, ultimately benefiting the well-being of older individuals through improved memory and a reduced risk of cognitive impairment.

## 5. Conclusion

This proof-of-concept study demonstrates that memory-related brainwave patterns and cognitive-motor functions in the aging brain can be modulated through neurofeedback training, even with a limited number of sessions. We developed a neurofeedback protocol to reinforce brainwave patterns associated with optimal working memory retrieval, and the preliminary results support our hypothesis: rewarding these brain patterns enhances cognitive efficiency and promotes more youthful brain oscillations. Specifically, post-training resting EEG measurements showed an increase in alpha oscillations and decreased in delta power, suggesting a reversal of age-related “neural slowing.” Furthermore, NF-rewarded trials displayed a more focused and efficient brain network compared to unrewarded trials. This memory-related NF protocol is a promising candidate for clinical trials as a non-pharmacological intervention to enhance cognitive function and reduce the risk of cognitive decline in aging individuals, as well as in patients with high-risk of mild cognitive impairment.

## Data Availability

All data produced in the present study are stored at the Lexington VA HCS and the University of Kentucky, which are available upon reasonable request to the corresponding author.

## Acknowledgements

The manuscript was partially supported by United States Department of Veterans Affairs Funding 5I21RX003173 and United States National Institute of Health (NIH), National Institute of Aging grant R56AG060608. The authors thank J. Suhl, MD and G. Markowski, BS for their inputs and assistance in data collection, and V. Zacharious for comments on an earlier version. The contents of this publication are solely the responsibility of the authors and do not represent the official views of these Institutions. The funders played no role in preparation of the manuscript or decision to publish.

